# Maternal postnatal depression and offspring depression at age 24 years in a UK-birth cohort: the mediating role of maternal nurturing behaviours concerning feeding, crying and sleeping

**DOI:** 10.1101/2020.06.22.20137331

**Authors:** Iryna Culpin, Gemma Hammerton, Marc H Bornstein, Jon Heron, Jonathan Evans, Tim Cadman, Hannah M Sallis, Kate Tilling, Alan Stein, Alex SF Kwong, Rebecca M Pearson

## Abstract

**Importance:** Few longitudinal studies have examined the role of maternal nurturing parenting behaviours concerning feeding, crying and sleeping in the association between maternal postnatal depression (PND) and offspring depression in adulthood.

**Objective:** To examine the association between PND and offspring depression at age 24 years and the mediating role of maternal nurturing behaviours concerning feeding, crying and sleeping.

**Design:** Longitudinal study of mothers and their offspring in the Avon Longitudinal Study of Parents and Children, followed up through age 24 years. Offspring ICD-10 depression diagnosis at age 24 years was established using the Clinical Interview Schedule-Revised. Symptoms of maternal depression were measured using the Edinburgh Postnatal Depression Scale at 8 months postnatally. Maternal nurturing behaviours concerning feeding, crying and sleeping were assessed using self-reported questionnaires administered from birth to age 3 years.

**Results:** The sample with complete data on confounders for the mediation analyses was 5,881. In the fully adjusted model, there was evidence of an indirect effect from PND to offspring depression through the combination of all parenting factors (probit regression coefficient [*B*]=0.038, 95%CI 0.005, 0.071); however, there was no evidence of a direct effect from early maternal PND to adult offspring depression once the indirect effect via parenting factors was accounted for (*B*=0.009, 95%CI -0.075, 0.093). There was evidence for specific indirect effects through maternal worries about feeding (*B*=0.019, 95%CI 0.003, 0.035, p=0.010) and maternal perceptions and responses to crying (*B*=0.018, 95%CI 0.004, 0.032, p=0.012). Analyses in a larger sample using multiple imputation led to similar results.

**Conclusions and Relevance:** The adverse impact of PND on offspring depression in early adulthood was explained by maternal nurturing behaviours concerning feeding, crying and sleeping in early infancy. Residual confounding and measurement error are likely, limiting causal conclusions. If found to be causal, reducing worries around early maternal nurturing behaviours could be a target for interventions to reduce adverse outcomes in adult offspring of depressed mothers.

## Introduction

Links between maternal postnatal depression (PND) and increased risk of later psychological problems in offspring are unquestioned.^1^ However, few large prospective longitudinal studies have examined whether these adverse effects persist into adulthood (18-20 years of age).^2^ Furthermore, the offspring outcomes associated with maternal PND are heterogenous and effect sizes are small to moderate.^1^ Thus, it is important to elucidate putative mediating factors that underly associations between parental and offspring depression. Such insights would be crucial to highlight those at greater risk and develop targeted interventions to reduce adverse outcomes in offspring of depressed mothers.

A substantial body of evidence suggests that an important potential mediator is the quality of parenting.^1, 3^ Specifically, PND disrupts maternal sensitivity^4^ and is associated with less engaged parenting,^5^ which is, in turn, associated with adverse offspring outcomes, including mental health problems.^6^

Numerous studies have identified aspects of parenting that may be reflective of the affective, cognitive and physical symptoms that characterise PND,^5^ including lower warmth, sensitivity and responsiveness.^6^ However, much less attention has been dedicated to the importance of the application of lower responsiveness and sensitivity to the everyday basic nurturing activities that are essential to infant care, or their potential explanatory role in the association between maternal and offspring depression. Caring for infants, who are fully dependent on their parents, requires the highest level of involvement often focused on meeting children’s basic needs such as feeding and managing crying and sleeping routines. Importantly, such basic needs are impossible to ignore, unpredictable and stressful, thus, may be particularly challenging for parents with depression. Infant crying peaks in the first 3 months, including an increase in prolonged night-time crying,^7^ while initiating and maintaining sleep is a common problem during the first years, challenging parents with long evening sleep rituals, waking at night and coming into parental bed.^8^ Approximately 20-25% of parents report problems feeding infants in the first 2 years,^9^ with food refusal and fussiness being a common feature of eating behaviour in young children.^10^ Although feeding, crying and sleeping patterns in young children are highly variable, they nevertheless are universal features of early child development, thus, calling for a range of parenting managements strategies.

Ample evidence established a link between maternal PND and sleep difficulty in infants and young children,^11,12^ including increased frequency of child awakenings.^13^ This may have an impact on how mothers approach bedtime routines, including regularity, sleep ecology (i.e., sleep location) and night waking behaviours. Similarly, PND impacts child feeding behaviour, including food refusal and fussy eating.^14^ PND may also impact maternal feeding behaviours (i.e., feeding style and practices), with depressed mothers using more physical and verbal pressure and offering more incentives to encourage their children to eat.^15^ PND has also been associated with more crying per day,^16^ longer episodes of crying/fussing and increased crying bout frequency,^17^ as well as reduced sensitivity and responsiveness to infant crying (e.g., feeding, rocking and touching).^18^

Parenting is part of a transactional dynamic^19^ with children shaping parenting behaviour.^20^ Children who are more difficult to look after due to frequent crying, fussy eating and poor sleep routines may evoke mood changes in caregivers and trigger more reactive and less consistent parenting.^1^ These so-called ‘evocative’ child effects are important to account for as findings regarding parental influences on child outcomes may partially reflect child influences.^20^ This may be of particular importance in the context of maternal and offspring depression, which share common genetic liabilities.^21^

Although quality of parenting may be important for offspring,^22^ little is known regarding the role that maternal basic nurturing parenting behaviours concerning feeding, crying and sleeping (henceforth referred to as maternal nurturing behaviours) play in the association between maternal and offspring depression. In the current study we address this gap by examining the impact of PND on such nurturing parenting behaviours and estimating the extent to which the association between PND and offspring depression at age 24 years is explained by early parenting using data from a large UK-based birth cohort study, the Avon Longitudinal Study of Parents and Children (ALSPAC). Quantifying this association may inform preventative and intervention programmes given that behavioural parenting strategies are modifiable.^23^ The richness of the ALSPAC data provides a unique opportunity to account for a number of factors associated with both maternal and offspring depression, including child polygenic score for neuroticism that may indicate genetic confounding. Our specific research questions were:

1. Is maternal PND associated with offspring depression at age 24 years?
2. Is any observed association mediated by maternal nurturing behaviours?

## Methods

### Study cohort

The sample comprised participants from the ALSPAC cohort. During Phase I enrolment, 14,541 pregnant mothers residing in the former Avon Health Authority in the south-west of England with expected dates of delivery between 1 April 1991 and 31 December 1992 were recruited. The total sample size is 15,247 pregnancies, of which 14,701 were alive at 1 year of age. Our sample comprised 12,986 mothers with at least one parenting item. Ethical approval and informed consent for the data collection was obtained from the ALSPAC Ethics and Law Committee and the Local Research Ethics Committees. Information about ALSPAC is available at www.bristol.ac.uk/alspac/, including a searchable data dictionary (http://www.bris.ac.uk/alspac/researchers/our-data). Further details on the cohort profile, representativeness and phases of recruitment are described in two cohort-profile papers.^24,25^

## Measures

### Exposure: maternal postnatal depression

Symptoms of maternal PND were measured using the Edinburgh Postnatal Depression Scale (EPDS),^26^ a 10-item self-reported depression questionnaire validated for use during the perinatal period and administered to mothers at 8 weeks postnatally. Confirmatory Factor Analysis (CFA) was used to derive a normally distributed latent trait based on 10-EPDS ordinal response items (see Methods S1, available online).

### Outcome: offspring depression

Offspring depression was assessed using the computerised version of the Clinical Interview Schedule-Revised (CIS-R),^27^ a fully structured psychiatric interview widely used in the community samples. It was administered at the research clinic at age 24 years to identify individuals with an ICD-10 diagnosis of depression (*versus* no diagnosis).

### Mediators: maternal nurturing behaviours

Full details pertaining to item selection and development of factors encapsulating maternal nurturing behaviours concerning feeding, crying and sleeping are presented in Methods S1 (available online). We extracted items from maternal self-reported questionnaires administered from birth to age 3 years 6 months (8 occasions) capturing maternal feeding style and practices, perceptions and responses to crying, and strategies to regulate bedtime routine and sleep ecology. These items were entered into separate CFA models per each dimension.

### Potential confounders: child polygenic score for neuroticism, socioeconomic, parental and family characteristics

Disadvantaged socioeconomic status and marital conflict are strong risk factors for perinatal depression^28^ and less optimal parenting practices.^29^ Thus, we adjusted for a range of potential confounding factors collected prospectively from maternal questionnaires during the antenatal period: highest maternal educational attainment (minimal education or none/compulsory secondary level (up to age 16 years; O-Level) *versus* non-compulsory secondary level (up to age 18 years; A-Level)/university level education), maternal age in years, family size (<3 *versus* ≥3 children), early parenthood (dichotomised as ≤19 years *versus* ≥ 20 years), perceived affordability of the cost of living (yes *versus* no), and parental conflict/aggression derived from questions asking how the mother and her current partner behaved towards each other (yes *versus* no). Analyses were also adjusted for child polygenic score (PGS) for neuroticism to account for possible genetic confounding.^1^ Genotyped data were available for 8,237 children in the ALSPAC (full details in Methods S1, available online).

## Statistical analysis

### Latent factor model

The hypothesised latent factor model is represented in Figure S1 (available online). Full details of latent factor model derivation, including the flow chart of items included into the CFA and derived factors and factor loadings are presented in Figure S2 and Table S1 (available online). In summary, items that were both theoretically relevant and showed standardised loadings (>0.15) on the relevant parenting dimension were included into a combined model using CFA with a robust Weighted Least Square (WLSMV) estimator to model categorical and continuous data.^30^ Root Mean Square Error of Approximation (RMSEA; >0.06), Comparative Fit Index (CFI) and Tucker-Lewis Index (TLI; >0.95) were used to evaluate model fit.^31^ The chi-square test of overall fit is prone to model misspecification when sample size is large,^32^ thus, we gave preference to relative fit indices.

### Direct and mediated effects

Multifaceted constructs, such as different aspects of parenting, are challenging and important in mediation as each specific factor may relate differentially to the outcome.^33^ Full details of the mediation model to examine direct and indirect (mediated) effects are presented in Method S1 (available online). In summary, we examined the extent to which the association between maternal postnatal (8 weeks) and offspring (24 years) depression (direct effect) is mediated by specific factors related to maternal nurturing behaviours (0-3 years; indirect effects). Analyses of longitudinal mediation models were restricted to those with complete data on the child neuroticism PGS and antenatal confounders (n=5,881). We used Structural Equation Modelling (SEM) in M*plus* v.8.2^34^ with latent constructs of maternal depression and parenting to estimated unadjusted (Model ^a^: including exposure, outcome and mediator only) and incrementally adjusted models (Model ^a+b^: adjusted for child neuroticism PGS; Model ^a+b+c^: further adjusted for socioeconomic and maternal characteristics; Model ^a+b+c+d^: further adjusted for parental conflict; Table 1). Results from path analyses with binary outcome (offspring diagnosis of depression), including indirect effects, are presented as probit regression coefficients (hereafter referred to as *B*, details in Methods S1). Probit coeffecients represent the predicted probability of the outcome for a 1 unit increase in the exposure (i.e., PND). Unadjusted and adjusted mediation models are presented in Figures 1-2. We conducted sensitivity analyses with early diagnosis of offspring depression (CIS-R)^27^ and depressive symptoms (MFQ)^35^ at 18 years (Results S1, available online).

**Table 1.**
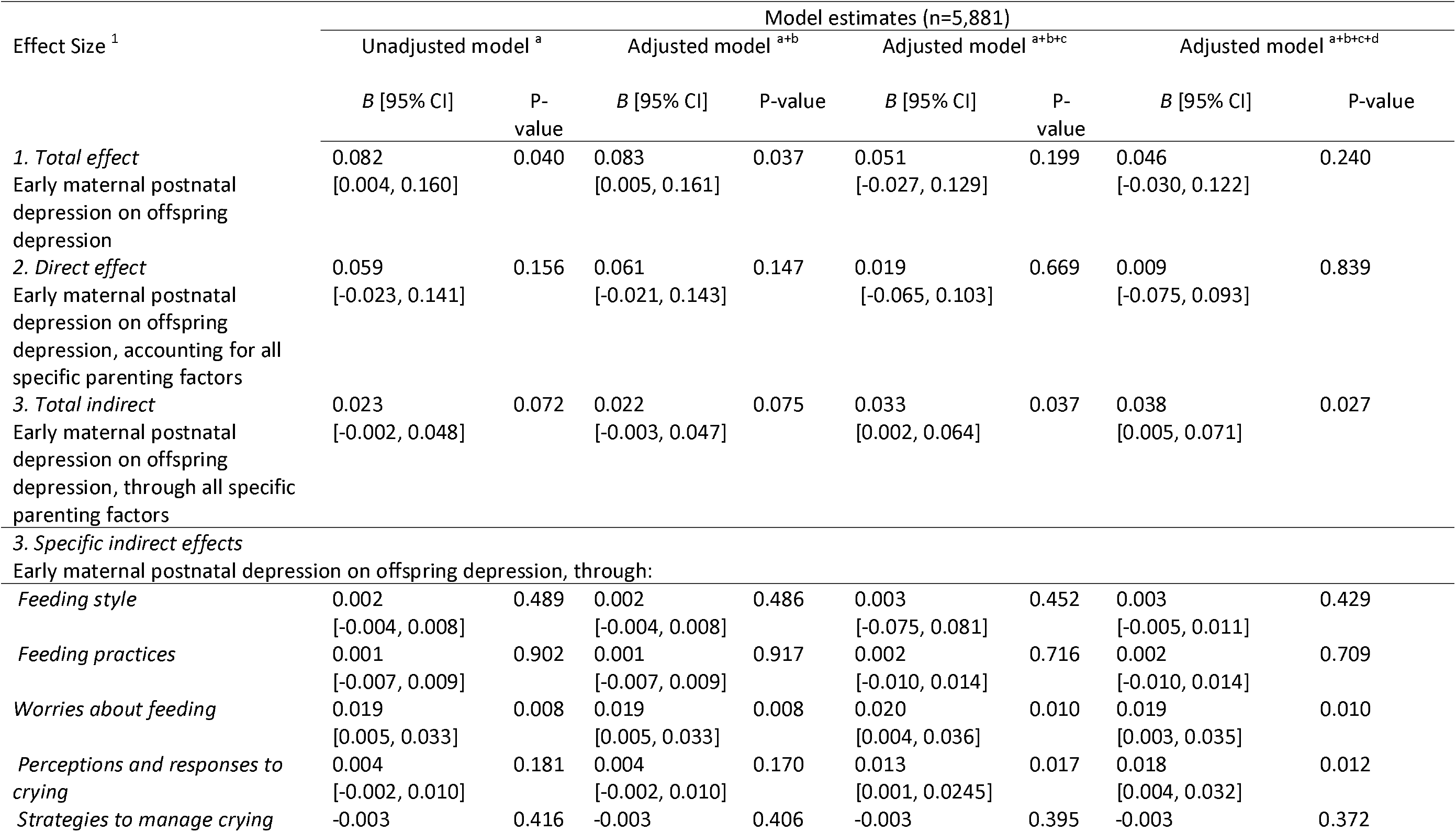
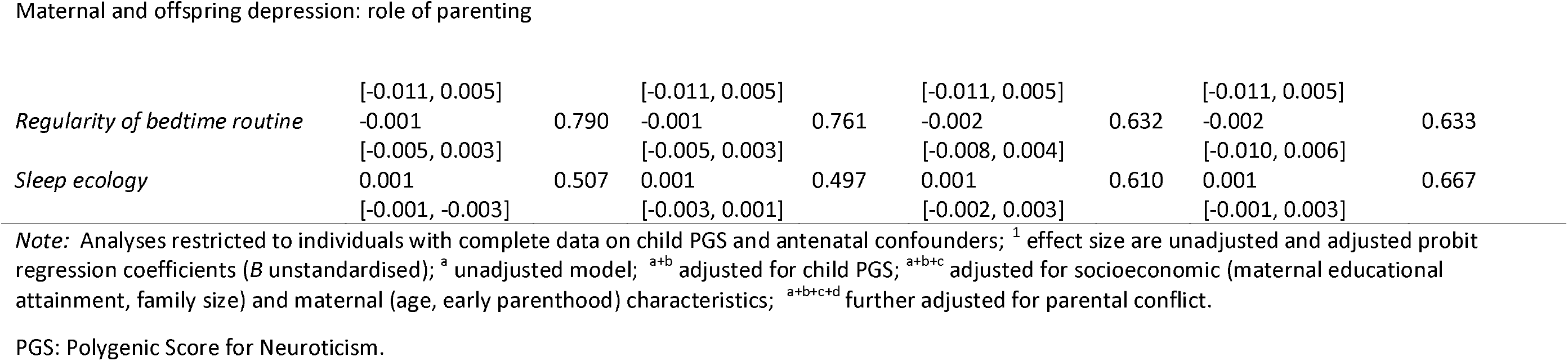
Estimates of the direct and mediated effects in the specific factors mediator model unadjusted and adjusted for child PGS and antenatal confounders in complete sample (n=5,881; exposure: maternal depression modelled as a latent factor)

| Effect Size <sup>1</sup> | Model estimates (n=5,881) |  |  |  |  |  |  |  |
| --- | --- | --- | --- | --- | --- | --- | --- | --- |
|  | Unadjusted model <sup>a</sup> |  | Adjusted model <sup>a+b</sup> |  | Adjusted model <sup>a+b+c</sup> |  | Adjusted model <sup>a+b+c+d</sup> |  |
|  | B [95% CI] | P-value | B [95% CI] | P-value | B [95% CI] | P-value | B [95% CI] | P-value |
| *1. Total effect* | 0.082 | 0.040 | 0.083 | 0.037 | 0.051 | 0.199 | 0.046 | 0.240 |
| Early maternal postnatal depression on offspring depression | [0.004, 0.160] |  | [0.005, 0.161] |  | [-0.027, 0.129] |  | [-0.030, 0.122] |  |
| *2. Direct effect* | 0.059 | 0.156 | 0.061 | 0.147 | 0.019 | 0.669 | 0.009 | 0.839 |
| Early maternal postnatal depression on offspring depression, accounting for all specific parenting factors | [-0.023, 0.141] |  | [-0.021, 0.143] |  | [-0.065, 0.103] |  | [-0.075, 0.093] |  |
| *3. Total indirect* | 0.023 | 0.072 | 0.022 | 0.075 | 0.033 | 0.037 | 0.038 | 0.027 |
| Early maternal postnatal depression on offspring depression, through all specific parenting factors | [-0.002, 0.048] |  | [-0.003, 0.047] |  | [0.002, 0.064] |  | [0.005, 0.071] |  |
| *3. Specific indirect effects* |  |  |  |  |  |  |  |  |
| Early maternal postnatal depression on offspring depression, through: |  |  |  |  |  |  |  |  |
| *Feeding style* | 0.002 | 0.489 | 0.002 | 0.486 | 0.003 | 0.452 | 0.003 | 0.429 |
|  | [-0.004, 0.008] |  | [-0.004, 0.008] |  | [-0.075, 0.081] |  | [-0.005, 0.011] |  |
| *Feeding practices* | 0.001 | 0.902 | 0.001 | 0.917 | 0.002 | 0.716 | 0.002 | 0.709 |
|  | [-0.007, 0.009] |  | [-0.007, 0.009] |  | [-0.010, 0.014] |  | [-0.010, 0.014] |  |
| *Worries about feeding* | 0.019 | 0.008 | 0.019 | 0.008 | 0.020 | 0.010 | 0.019 | 0.010 |
|  | [0.005, 0.033] |  | [0.005, 0.033] |  | [0.004, 0.036] |  | [0.003, 0.035] |  |
| *Perceptions and responses to crying* | 0.004 | 0.181 | 0.004 | 0.170 | 0.013 | 0.017 | 0.018 | 0.012 |
|  | [-0.002, 0.010] |  | [-0.002, 0.010] |  | [0.001, 0.0245] |  | [0.004, 0.032] |  |
| *Strategies to manage crying* | -0.003 | 0.416 | -0.003 | 0.406 | -0.003 | 0.395 | -0.003 | 0.372 |
|  | [-0.011, 0.005] |  | [-0.011, 0.005] |  | [-0.011, 0.005] |  | [-0.011, 0.005] |  |
| *Regularity of bedtime routine* | -0.001 | 0.790 | -0.001 | 0.761 | -0.002 | 0.632 | -0.002 | 0.633 |
|  | [-0.005, 0.003] |  | [-0.005, 0.003] |  | [-0.008, 0.004] |  | [-0.010, 0.006] |  |
| *Sleep ecology* | 0.001 | 0.507 | 0.001 | 0.497 | 0.001 | 0.610 | 0.001 | 0.667 |
|  | [-0.001, -0.003] |  | [-0.003, 0.001] |  | [-0.002, 0.003] |  | [-0.001, 0.003] |  |

*Note:* Analyses restricted to individuals with complete data on child PGS and antenatal confounders; <sup>1</sup> effect size are unadjusted and adjusted probit regression coefficients (*B* unstandardised); <sup>a</sup> unadjusted model; <sup>a+b</sup> adjusted for child PGS; <sup>a+b+c</sup> adjusted for socioeconomic (maternal educational attainment, family size) and maternal (age, early parenthood) characteristics; <sup>a+b+c+d</sup> further adjusted for parental conflict.
PGS: Polygenic Score for Neuroticism.

**Figure 1.**
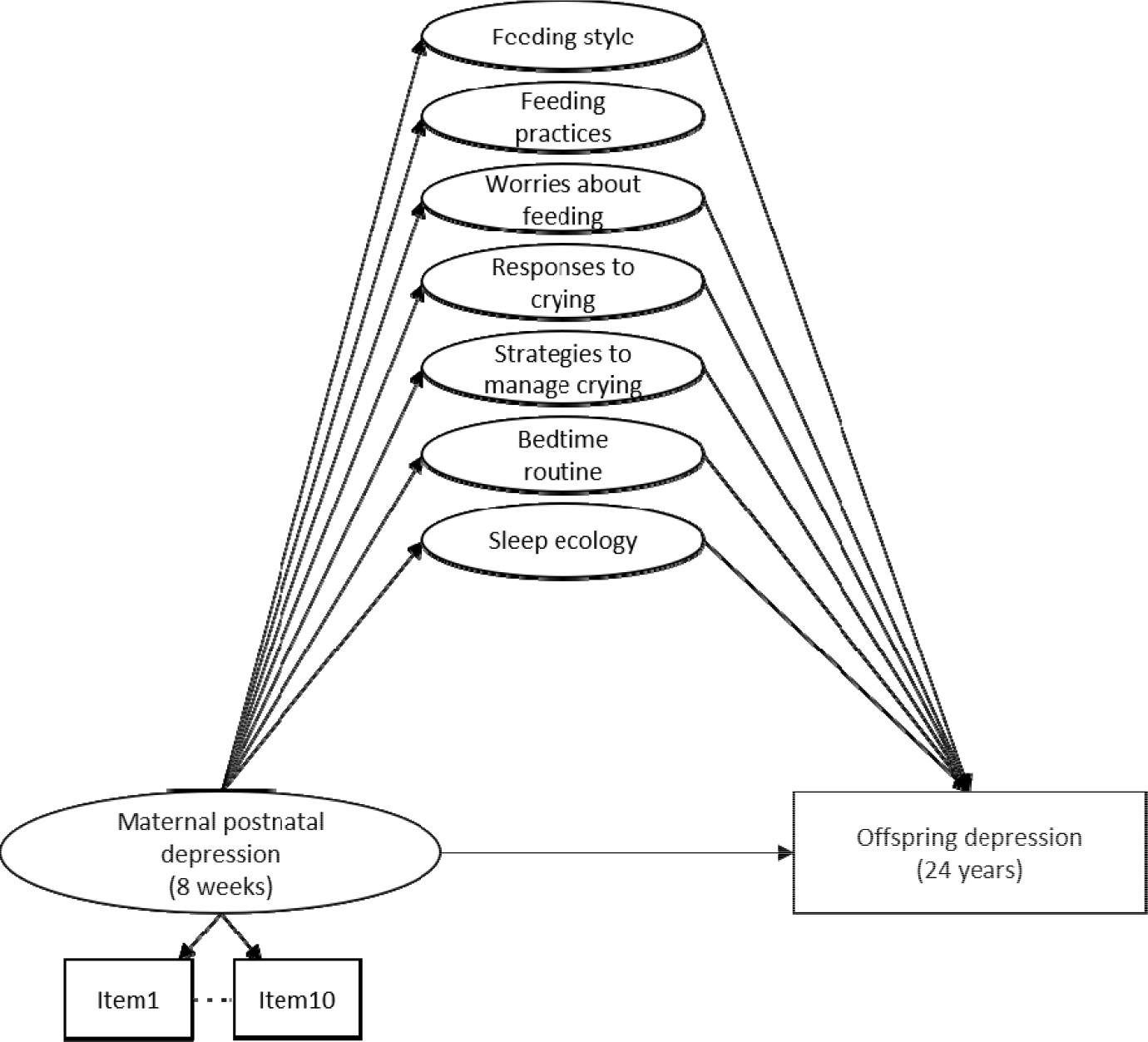
Hypothesised specific factors mediation model (unadjusted) Note: Observed items and variables are represented by squares, whilst latent variables are represented by circles. Error terms covariances and individual items loading onto each specific factor are not represented to reduce figure complexity.

**Figure 2.**
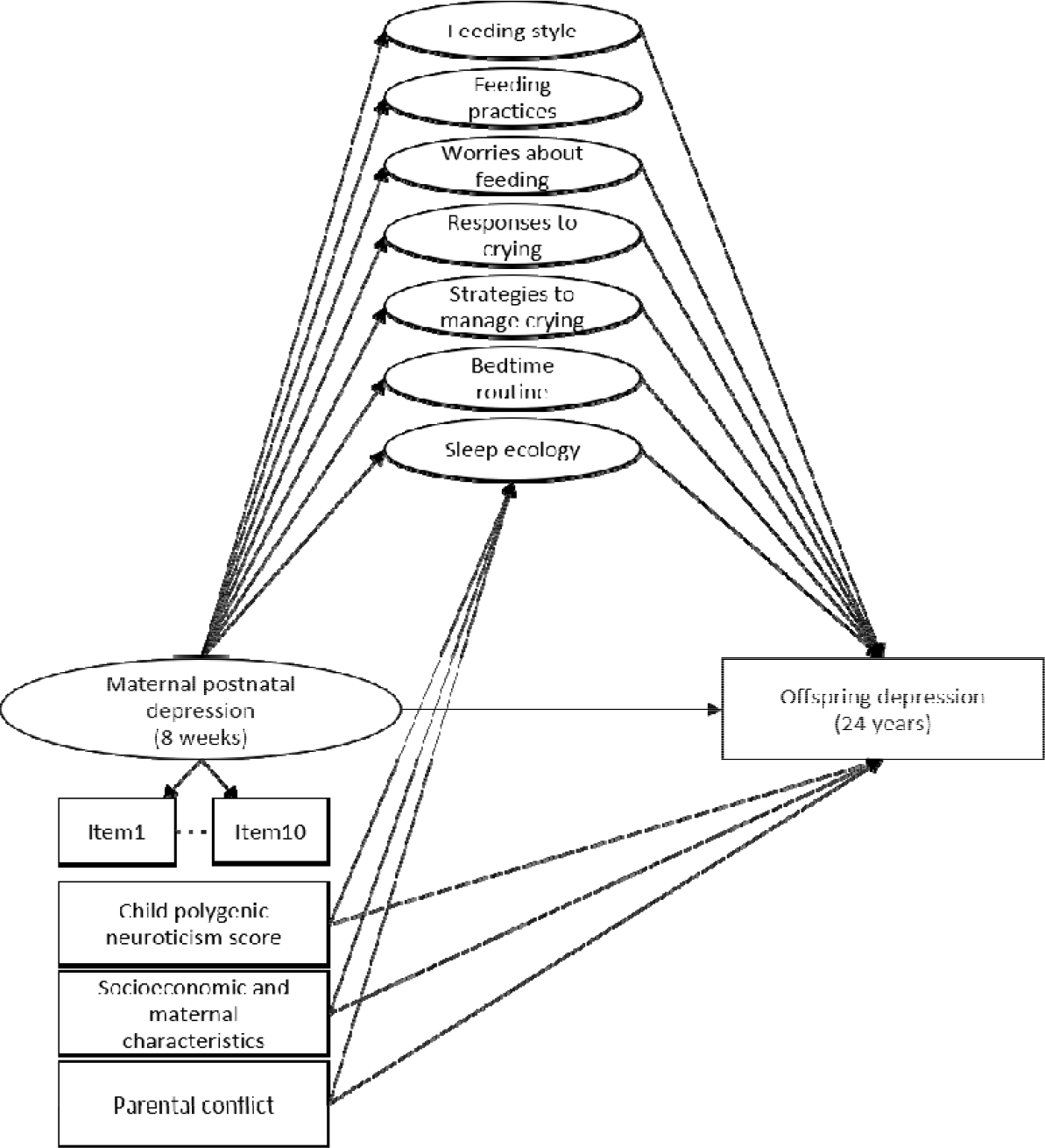
Structural specific factors mediation model estimating the direct effect of maternal early postnatal depression on offspring depression at age 24 years and the indirect effect through seven specific crying, feeding and sleeping maternal parenting factors, adjusted for child polygenic neuroticism score, socioeconomic (maternal educational attainment, family size) and maternal (age, early parenthood) characteristics, and parental conflict Note: Observed variables are represented by squares, whilst the latent variable is represented by a circle. Error terms covariances and individual items loading onto each specific factor are not represented to reduce figure complexity. All specific factors in the model were allowed to correlate.

### Missing data: multiple imputation

We conducted sensitivity analyses to examine the impact of missing data on our findings. Full description of the imputation method is presented in Method S1 (available online).

## Results

The characteristics of our study sample and prevalence of offspring depression at age 24 years by the presence of maternal PND are presented in Table S2 (available online). There were relatively high correlations between feeding, crying and sleep-related maternal parenting behaviours (Results S1; Table S3, available online). The associations between PND and maternal nurturing parenting behaviours, as well as parenting behaviours and offspring depression are described in Results S1 and Table S4. In summary, maternal PND was strongly associated with less optimal maternal nurturing behaviours. Maternal worries regarding child’s feeding and less sensitive perceptions and responses to crying were associated with higher risk of offspring depression at age 24 years.

### Factors encapsulating maternal nurturing behaviours

A model using CFA to fit latent factors capturing maternal nurturing behaviours suggested an adequate measurement model fit (RMSEA: 0.033, 95%CI 0.032 to 0.033; CFI/TLI: 0.926/0.918; Table 1S, available online) supporting further tests of structural paths (direct and mediated effects). Seven factors representing maternal nurturing behaviours concerning feeding, crying and sleeping were derived (full details in Results S1, available online).

*Factor 1 Feeding style*: maternal overall attitude regarding feeding, with higher factor scores representing maternal feeding style more concordant with authoritative parenting (i.e., higher levels of maternal responsiveness and appreciation of feeding routine).

*Factor 2 Feeding practices*: maternal approaches to feeding, with higher factor scores representing less pressured and restricting feeding behaviour.

*Factor 3 Worries about feeding*: maternal worries regarding child’s feeding, with higher factor scores representing higher levels of maternal worry regarding child’s feeding, so unlike other factors higher scores are predicted to confer greater risk.

*Factor 4 Perceptions of and responses to crying*: maternal feelings and behaviours in response to child’s crying, with higher factor scores representing more sensitive maternal responses to child’s crying.

*Factor 5 Strategies to manage crying*: maternal strategies to deal with child’s crying, with higher factor scores representing more optimal responses to child’s crying

*Factor 6 Regularity of bedtime routine*: maternal behaviours to regulate bedtime routine, with higher factor scores representing more maternal adherence to regular sleep routine and less reactive response to child’s non-compliance with bedtime routine.

*Factor 7 Sleep ecology*: maternal strategies to manage child’s sleep location and night walking behaviours, with higher factor scores representing more consistent and adaptive maternal responses to child’s night walking behaviours.

### Direct and mediated effects

We estimated unadjusted and adjusted structural mediation models to examine the direct effect of maternal PND on offspring depression and the mediated effects through specific parenting factors whilst accounting for a range of confounders. Analyses were restricted to those with complete data on the child neuroticism PGS and antenatal confounders (n=5,881). Of 3,567 young adults with data on depression diagnosis at age 24 years, 384 had depression (10.8%, 95%CI 0.09, 0.12). There was some evidence of total indirect effect from early maternal PND to offspring depression at age 24 years through the combination of all specific parenting factors in the unadjusted (probit regression coefficient [*B*] =0.023, 95%CI -0.002, 0.048, p=0.072) and fully adjusted models (*B* =0.038, 95%CI 0.005, 0.071, p=0.027), although the 95% CIs were wide. There was no evidence of a direct effect from maternal PND to offspring depression at age 24 years in the unadjusted (*B* =0.059, 95%CI - 0.023, 0.141, p=0.156) and fully adjusted (*B* =0.009, 95%CI -0.075, 0.093, p=0.839) models once the indirect effect via parenting factors was accounted for (Table 1). There was some evidence for specific indirect effects through maternal worries about feeding (*B* =0.019, 95%CI 0.003, 0.035, p=0.010) and maternal perceptions and responses to crying in the fully adjusted models (*B* =0.018, 95%CI 0.004, 0.032, p=0.012). Sensitivity analyses using bias-corrected (BC) bootstrapping (n=1,000)^36^ to estimate indirect effect led to similar conclusions, albeit with higher p-values (Table S5, available online).

It was only possible to model the EPDS as a sum-score in imputed data analyses due to rare values on specific individual items. Thus, to investigate the impact of missing data we compared equivalent models in complete (n=5,881) and imputed (n=7,523) data sets using the EPDS as a sum-score. The results from the analyses with imputed data (Table 2; Results 1, available online) supported our findings: the total indirect effects were in the same direction and led to the same over-arching conclusions as in the complete case analyses using EPDS as a sum-score (Table 3). However, there was stronger evidence for the total indirect effect in the imputed data analyses (Table 2; unadjusted model: *B*=0.007, 95%CI 0.001, 0.013, p=0.012 ; fully adjusted model: *B*=0.005, 95%CI 0.001, 0.009, p=0.026) compared to complete case analyses (Table 3: unadjusted model: *B*=0.007, 95%CI -0.001, 0.015, p=0.088; fully adjusted model: *B*=0.006, 95%CI -0.002, 0.015, p=0.071). The sensitivity analyses with diagnosis of depression (CIS-R)^27^ and depressive symptoms (MFQ)^35^ at 18 years resulted in the same pattern of results, with stronger evidence for the direct effect in the fully adjusted model with depressive symptoms as an outcome (Results S1 and Table S6, available online).

**Table 2.**
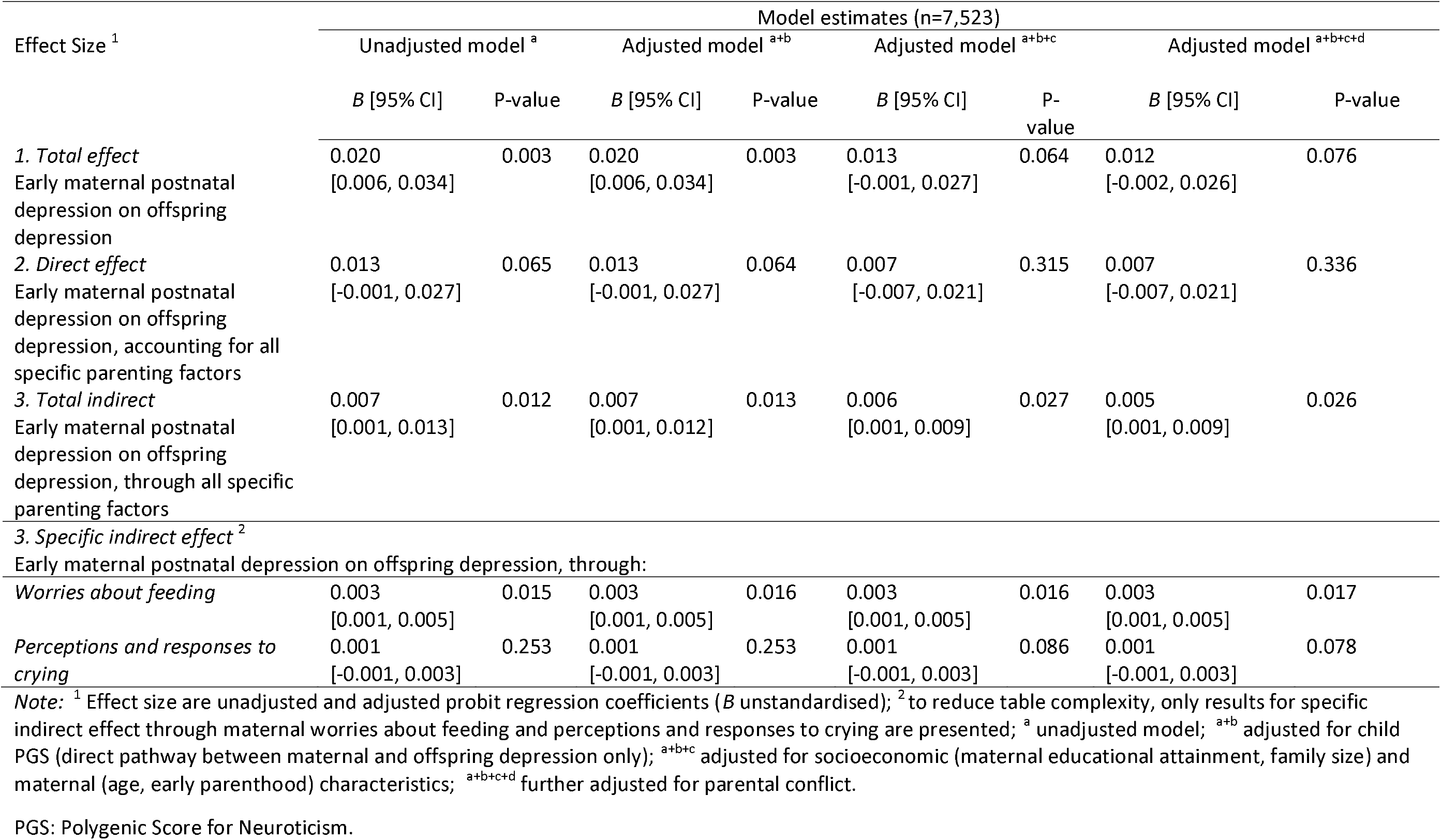
Estimates of the direct and mediated effects in the specific factors mediator model unadjusted and adjusted for child PGS and antenatal confounders in imputed sample (n=7,523; exposure: maternal depression modelled as a sum-score)

**Table 3.** Estimates of the direct and mediated effects in the specific factors mediator model unadjusted and adjusted for child PGS and antenatal confounders in complete sample (n=5,881; exposure: maternal depression modelled as a sum-score)

| Effect Size <sup>1</sup> | Model estimates (n=5,881) |  |  |  |  |  |  |  |
| --- | --- | --- | --- | --- | --- | --- | --- | --- |
|  | Unadjusted model <sup>a</sup> |  | Adjusted model <sup>a+b</sup> |  | Adjusted model <sup>a+b+c</sup> |  | Adjusted model <sup>a+b+c+d</sup> |  |
|  | B [95% CI] | P-value | B [95% CI] | P-value | B [95% CI] | P-value | B [95% CI] | P-value |
| 1. Total effect | 0.016 | 0.047 | 0.016 | 0.044 | 0.010 | 0.218 | 0.009 | 0.257 |
| Early maternal postnatal depression on offspring depression | [0.001, 0.032] |  | [0.001, 0.032] |  | [-0.006, 0.026] |  | [-0.007, 0.025] |  |
| 2. Direct effect | 0.009 | 0.306 | 0.009 | 0.287 | 0.004 | 0.678 | 0.003 | 0.736 |
| Early maternal postnatal depression on offspring depression, accounting for all specific parenting factors | [-0.009, 0.027] |  | [-0.009, 0.027] |  | [-0.013, 0.022] |  | [-0.015, 0.021] |  |
| 3. Total indirect | 0.007 | 0.088 | 0.007 | 0.044 | 0.006 | 0.080 | 0.006 | 0.071 |
| Early maternal postnatal depression on offspring depression, through all specific parenting factors | [-0.001, 0.015] |  | [-0.001, 0.015] |  | [-0.002, 0.014] |  | [-0.002, 0.014] |  |
| 3. Specific indirect effect <sup>2</sup> |  |  |  |  |  |  |  |  |
| Early maternal postnatal depression on offspring depression, through: |  |  |  |  |  |  |  |  |
| Worries about feeding | 0.004 | 0.009 | 0.004 | 0.010 | 0.004 | 0.011 | 0.004 | 0.012 |
|  | [0.001, 0.008] |  | [0.001, 0.008] |  | [0.001, 0.008] |  | [0.001, 0.008] |  |
| Perceptions and responses to crying | 0.001 | 0.073 | 0.001 | 0.074 | 0.002 | 0.030 | 0.002 | 0.027 |
|  | [-0.001, 0.003] |  | [-0.001, 0.003] |  | [0.001, 0.004] |  | [0.001, 0.004] |  |

Note: <sup>1</sup> Effect size are unadjusted and adjusted probit regression coefficients (*B* unstandardised); <sup>2</sup> to reduce table complexity, only results for specific indirect effect through maternal worries about feeding and perceptions and responses to crying are presented; <sup>a</sup> unadjusted model; <sup>a+b</sup> adjusted for child neuroticism score (direct pathway between maternal and offspring depression only); <sup>a+b+c</sup> adjusted for socioeconomic (maternal educational attainment, family size) and maternal (age, early parenthood) characteristics; <sup>a+b+c+d</sup> further adjusted for parental conflict.
PGS: Polygenic Score for Neuroticism.

## Discussion

In this population-based cohort study, we found evidence for an association between maternal PND and increased risk of offspring depression at age 24 years, which was largely explained by maternal nurturing behaviours during early childhood. Maternal PND was associated with less maternal nurturing behaviours, which, in turn, were associated with increased risk of offspring depression. Indeed, in the fully adjusted model, there was robust evidence of an indirect pathway through maternal parenting behaviours, but no evidence of a remaining direct association between maternal and offspring depression. The indirect pathway was driven by two specific parenting factors, maternal worries about feeding and perceptions and responses to crying.

Our findings are consistent with previous longitudinal research linking parental depression with less optimal parenting behaviour,^37^ which in turn increases the risk for offspring depression.^38,39^ Bifulco et al.^40^ found that maternal depression had no direct effect on offspring psychopathology, once a mediating pathway through a composite measure of offspring neglect and abuse was accounted for. However, these studies focused on harsh dimensions of parenting,^39,40^ with less emphasis on day-to-day parenting strategies to meet and attend to infants’ basic nurturing needs. This study extends existing literature with evidence that variance in these nurturing parenting domains is an important explanatory mechanism in the association between maternal and offspring depression. The negative emotions that characterise maternal depression may be colouring the experience of day-to-day parenting and maternal responses to feeding and crying.^41^

We found evidence of specificity for two parenting factors in mediating the mother-to-child depression effect, namely maternal worries about feeding and perceptions and responses to crying. Items around worries about feeding and crying were self-reported, which may not be reflective of actual maternal behaviours. However, what is shared between these two factors is emotional concern, which may be a critical component for the mediating mechanisms in the association between maternal and offspring depression. Mothers who experience depression are more prone to emotional dysregulation,^42^ which may translate into inconsistent or harsh parenting. Parent and child co-regulate emotions in infancy,^43^ thus, if the mother is feeling negative emotions when dealing with basic nurturing needs, the infant may also experience negative emotions. Consistent responsiveness to the infants’ needs also provides a predictable scaffolding, which empowers the infant to feel in control of their environment.^44^ However, where consistent responsiveness is disrupted, child’s self-regulatory competence may also be affected.

Associations between parenting and infant feeding, crying and sleeping are complex and, most likely, bidirectional.^19,20^ Our findings suggest that maternal PND is associated with maternal nurturing behaviours in early infancy. However, in line with transactional developmental models, infants with more difficult feeding, crying and sleeping patterns may influence maternal PND.^45^ Examination of possible bidirectionality was outside the scope of the current study. However, we accounted for possible evocative child effects and shared genetic liability for depression in mothers and offspring and related phenotypes such as parenting experiences by including genetic liability scores for neuroticism^46^. We only included genetic scores for neuroticism, which explained a small proportion of variance in that trait; thus shared genetic variance in depression and parenting not captured by such scores is likely to play a role in the associations between PND, parenting and offspring depression.

The strengths of the study include a longitudinal design and a large community-based sample that enabled us to examine the long-term association between maternal PND and offspring depression in early adulthood, as well as elucidate possible transmission pathways. To our knowledge, no previous studies have examined maternal nurturing behaviours as possible explanatory mechanisms in the association between maternal and offspring depression. Furthermore, we utilised clinical diagnosis of offspring depression and accounted for a range of confounders, including child neuroticism PGS. Modelling basic maternal nurturing behaviours concerning feeding, crying and sleeping as a latent factor enabled us to capture maternal behaviours across early childhood (birth to 3 years).

A limitation of the study relates to sample attrition, which is similar to that observed in other population-based studies.^24,25^ Sample attrition may have implications for internal validity, given that participants from lower socio-economic background and those with depression were under-represented in our complete sample. We addressed bias associated with selective attrition by controlling for factors known to be predictive of missingness and by imputing missing data in our exposure, outcome and confounders. The pattern of missing data and imputed analyses suggested that attrition may have led to an underestimation of the direct and indirect effects.

Non-independence of measurement and reporting bias, whereby maternal depression and parenting practices are reported by the same informant (i.e. the mother), is another limitation. Evidence suggests that depressed mothers may report more negative parenting,^47^ potential leading to the over-estimation of mediated effects. However, reports of specific behaviours assessed using relatively neutral/functional items (e.g., ‘frequency child made to go to bed’) may be less susceptible to bias than global assessment of parenting style.^42^ Even though maternal depression may influence reports of perceived worries surrounding parenting, the outcome in our study (offspring depression) was child-reported, suggesting that, at a minimum, maternal reports of their parenting are more predictive of offspring outcomes than reports of depression itself.

Our measure of parenting was self-reported rather than independently assessed, potentially biasing the estimation of associations between some parenting behaviours and offspring depression.^48^ However, we modelled parenting items across several time points, arguably capturing a more comprehensive picture of maternal behaviour compared to a one-off assessment. Although we adjusted for a range of possible measured confounders, residual confounding remains a possibility. The sizes of the associations were relatively small; however, in this study we looked at variance in maternal depression and parenting in the *whole* population sample where even small effects can have a meaningful impact.^49^ Finally, we didn’t explore measures of the fathers behaviour, which is known to be important.

Our findings indicate that maternal nurturing behaviours play an important role in the association between maternal and offspring depression. Further studies to examine whether these associations are causal are needed to strengthen these findings. If they are causal, interventions that identify and treat depression early, as well as enhance nurturing responses around parenting behaviours concerning feeding, crying and sleeping to address worries and emotional reactivity around such activities, may contribute toward reducing intergenerational transmission of mental health risk. This may be particularly important in light of recent evidence suggesting that parenting interventions focused on active acquisition of parenting skills and increased parenting confidence are effective in improving offspring development.^50^

## Ethical standards

The authors assert that all procedures contributing to this work comply with the ethical standards of the relevant national and institutional committees on human experimentation and with the Helsinki Declaration of 1975, as revised in 2008.

## Data Availability

Data available upon application for access submitted to ALSPAC Executive Committee.

## Acknowledgements

We are extremely grateful to all the families who took part in this study, the midwives for their help in recruiting them, and the whole ALSPAC team, which includes interviewers, computer and laboratory technicians, clerical workers, research scientists, volunteers, managers, receptionists and nurses. The UK Medical Research Council and Wellcome Trust (Grant ref: 102215/2/13/2) and the University of Bristol provide core support for ALSPAC. A comprehensive list of grants funding is available on the ALSPAC website (http://www.bristol.ac.uk/alspac/external/documents/grant-acknowledgements.pdf). This research was specifically funded by the European Research Commission awarded to Dr Pearson (Grant ref: 758813 MHINT). Dr Culpin is supported by the Wellcome Trust Research Fellowship in Humanities and Social Science (Grant ref: 212664/Z/18/Z). Dr Hammerton is supported by the Sir Henry Wellcome Postdoctoral Fellowship (209138/Z/17/Z). Professor Bornstein was funded by the Intramural Research Program of the NIH/NICHD, USA, and an International Research Fellowship at the Institute for Fiscal Studies (IFS), London, UK, funded by the European Research Council (ERC) under the Horizon 2020 research and innovation programme (grant agreement No: 695300-HKADeC-ERC-2015-AdG). Professor Stein was supported by the NIHR Oxford Health Biomedical Research Centre. Dr Cadman received funding from the European Union’s Horizon 2020 research and innovation programme under grant agreement N: 733206, LIFE-CYCLE project. Dr Sallis is a member of the MRC Integrative Epidemiology Unit at the University of Bristol (MC_UU_00011/7), which is supported by the UK Medical Research Council Unit (MC_UU_12013/3 and MC_UU_12013/4). This study was also supported by the NIHR Biomedical Research Centre at the University Hospitals Bristol NHS Foundation Trust and the University of Bristol. This publication is the work of the authors who will serve as guarantors for the contents of this paper. The views expressed in this publication are those of the author(s) and not necessarily those of the NHS, the National Institute for Health Research.

## Notes

### Competing Interest Statement

The authors have declared no competing interest.

### Clinical Trial

The study reported in the manuscript is based on a secondary analyses of a population-based birth cohort in Bristol (United Kingdom), not clinical trial.

### Author Declarations

Ethical approval and informed consent for the data collection was obtained from the ALSPAC Ethics and Law Committee and the Local Research Ethics Committees.

## References

1. Stein A, Pearson RM, Goodman SH, Rapa E, et al. Effects of perinatal mental disorders on the fetus and child. The Lancet. 2014;384:1800–1819.

2. Weissman MM, Berry OO, Warner V, et al. A 30-year study of 3 generations at high risk and low risk for depression. JAMA Psychiatry. 2016;73:970–977.

3. Sanger C, Iles JE, Andrew CS, Ramchandani PG. Associations between postnatal maternal depression and psychological outcomes in adolescent offspring: a systematic review. Arch Women Ment Hlth. 2015;18:147–162.

4. Murray L, Halligan SL, Cooper P. Effects of postnatal depression on mother-infant interactions, and child development. In: Handbook of infant development, eds. Malden, UK: Wiley-Blackwell; 2010.

5. Lovejoy MC, Graczyk PA, O’Hare E, Neuman G. Maternal depression and parenting behavior: a meta-analytic review. Clin Psychol Rev. 2000;20: 561–592.

6. Bornstein MH. Children’s Parents. In: Ecological settings and processes in developmental systems. Handbook of child psychology and developmental science. Hoboken, NJ: Wiley; 2015.

7. Walker AM, Menahem S. Normal early infant behaviour patterns. J Paediatr Child Health. 1994;30:260–262.

8. Wolke D. Frequent problems in the infancy and toddler years: excessive crying, sleeping, and feeding difficulties. In: Health promotion and disease prevention in the family. Berlin: Walter de Gruyter; 2003.

9. Lindberg L, Bohlin G, Hagekull B. Early feeding problems in a normal population. Int J Eat Disorder. 1991;10: 395–405.

10. Young B, Drewett R. Eating behaviour and its variability in 1-year-old children. Appetite. 2000;35:171–177.

11. Karraker KH, Young M. Night waking in 6-month-old infants and maternal depressive symptoms. J Appl Dev Psychol. 2007;28:493–498.

12. Stoleru S, Nottelmann ED, Belmont B, Ronsaville D. Sleep problems in children of affectively ill mothers. J Child Psychol Psychiatry. 1997;38:831–841.

13. Warren SL, Howe G, Simmens SJ, Dahl RE. Maternal depressive symptoms and child sleep: models of mutual influence over time. Dev Psychopathol. 2006;18:1–16

14. Coulthard H, Harris G. Early food refusal: the role of maternal mood. Journal of reproductive and infant psychology. 2003;21:335–345.

15. Haycraft E, Farrow C, Blissett J. Maternal symptoms of depression are related to observations of controlling feeding practices in mothers of young children. J Fam Psychol. 2013;27:159–164.

16. Milgrom J, Westley DT, McCloud PI. Do infants of depressed mothers cry more than other infants? J Paediatr Child Health. 1995;31:218–221.

17. Miller AR, Barr RG, Eaton WO. Crying and motor behavior of six-week-old infants and postpartum maternal mood. Pediatrics. 1993;92:551–558.

18. Esposito G, Manian N, Truzzi A, Bornstein MH. Response to infant cry in clinically depressed and non-depressed mothers. PloS One. 2017;12:e0169066.

19. Mills-Koonce WR, Propper CB, Gariepy JL, Blair C, Garrett-Peters P, Cox MJ. Bidirectional genetic and environmental influences on mother and child behavior: the family system as the unit of analyses. Dev Psychopathol. 2007;19:1073–1087.

20. Avinun, R., & Knafo, A. Parenting as a reaction evoked by children’s genotype: a meta-analysis of children-as-twins studies. Pers Soc Psychol Rev. 2014;18: 87–102.

21. Knafo, A., & Jaffee, S. R. Gene-environment correlation in developmental psychopathology. Dev Psych. 2013;25:1–6.

22. Smith, M. Good parenting: making a difference. Early Hum Dev. 2010;86:689–693.

23. Kaminski JW, Valle LA, Filene JH, Boyle CL. A meta-analytic review of components associated with parent training program effectiveness. J Abnorm Child Psych. 2008;36:567–589.

24. Boyd A, Golding J, Macleod J, et al. Cohort Profile: The ‘Children of the 90s’: the index offspring of the Avon Longitudinal Study of Parents and Children. Int J Epidemiol. 2013; 42:111–127.

25. Fraser A, Macdonald-Wallis C, Tilling K, et al. Cohort profile: the Avon Longitudinal Study of Parents and Children: ALSPAC mothers cohort. Int J Epidemiol. 2012:97–110.

26. Cox JL, Holden JM, Sagovsky R. Detection of postnatal depression: development of the 10-item Edinburgh Postnatal Depression Scale. Br J Psychiatry. 1987;150:782–786.

27. Lewis G, Pelosi AJ, Araya R, Dunn G. Measuring psychiatric disorder in the community: a standardized assessment for use by lay interviewers. Psychol Med. 1992;22:465–486.

28. Leigh B, Milgrom J. Risk factors for antenatal depression, postnatal depression and parenting stress. BMC Psychiatry. 2008;8:24.

29. Hoff E, Laursen B, Tardif T. Socioeconomic status and parenting. Handbook of Parenting: Biology and Ecology of Parenting. 2002;8:231–252.

30. Brown TA, Moore MT. Confirmatory factor analysis. In: Hoyle RH Handbook of structural equation modelling, eds. NY: Guilford Press; 2012.

31. Hu LT, Bentler PM. Fit indices in covariance structure modelling: Sensitivity to underparameterized model misspecification. Psychol Methods. 1998;3:424–453.

32. Lomax RG, Schumacker RE. A beginner’s guide to structural equation modeling. Psychology Press; 2004.

33. Gonzalez O, MacKinnon DP. A bifactor approach to model multifaceted constructs in statistical mediation analysis. Educ Psychol Meas. 2018;78:5–31.

34. Muthén LK, Muthén BO. Mplus User’s Guide, 7^th^ ed. Muthén & Muthén:Los Angeles, CA;2015.

35. Messer SC, Angold A, Costello EJ, Loeber R. Development of a short questionnaire for use in epidemiological studies of depression in children and adolescents: factor composition and structure across development. Int Jo Meth Psych Res. 1995;5:237–249.

36. MacKinnon DP, Lockwood CM, Williams J. Confidence limits for the indirect effect: distribution of the product and resampling methods. Multiva Behav Res. 2004;39:99–128.

37. Suveg C, Shaffer A, Morelen D, Thomassin K. Links between maternal and child psychopathology symptoms: mediation through child emotion regulation and moderation through maternal behavior. Child Psychiat Hum D. 2011;42:507–520.

38. Caron A, Weiss B, Harris Catron T. Parenting behavior dimensions and child psychopathology: specificity, task dependency, and interactive relations. J Clin Child Adolesc. 2006;35:34–45.

39. Johnson JG, Cohen P, Kasen S, Smailes E, Brook JS. Association of maladaptive parental behavior with psychiatric disorder among parents and their offspring. Arch Gen Psychiat. 2001;58:453–60.

40. Bifulco A, Moran PM, Ball C, et al. Childhood adversity, parental vulnerability and disorder: examining inter-generational transmission of risk. J Child Psychol Psyc. 2002;43:1075–1086.

41. Berg-Nielsen TS, Vikan A, Dahl AA. Parenting related to child and parental psychopathology: a descriptive review of the literature. Clin Child Psychol P. 2002;7:529–552.

42. Goodman SH, Gotlib IH. Risk for psychopathology in the children of depressed mothers: a developmental model for understanding mechanisms of transmission. Psychol Rev. 1999;106:458–490.

43. Calkins, SD. Caregiving as coregulation: Psychobiological processes and child functioning. In: Biosocial foundations of family processes. Springer, New York, NY; 2011.

44. Crittenden PM, Landini A. Attachment relationships as semiotic scaffolding systems. Biosemiotics. 2015;8:257–273.

45. Murray L, Stanley C, Hooper R, King F, Fiori-Cowley A. The role of infant factors in postnatal depression and mother-infant interactions. Dev Med Child Neurol. 1996;38:109–19.

46. Knafo A, Jaffee SR. Gene–environment correlation in developmental psychopathology. Dev Psychopathol. 2013;25:1–6.

47. Burt KB, Van Dulmen MH, Carlivati J, et al. Mediating links between maternal depression and offspring psychopathology: the importance of independent data. J Child Psychol Psyc. 2005;46:490–499.

48. Kendler KS, Baker JH. Genetic influences on measures of the environment: a systematic review. Psychol Med. 2007;37:615–26.

49. Bornstein MH. Human infancy … and the rest of the lifespan. Annu Rev Psychol. 2014;65:121–158.

50. Kaminski JW, Valle LA, Filene JH, Boyle CL. A meta-analytic review of components associated with parent training program effectiveness. J Abnorm Psychol. 2008;36:567–589.

